# Epidemiological health assessment in primary health care in the State of Qatar-2019

**DOI:** 10.1101/2021.02.27.21251797

**Authors:** Mohamed Ghaith Al-Kuwari, Samya Ahmad Al – Abdulla, Maha Yousef Abdulla, Ahmad Haj Bakri, Azza Mustafa Mohammed, Mujeeb Chettiyam Kandy, Amanda Patterson

## Abstract

**Objectives:** The Primary Health Care Corporation (PHCC) in Qatar conducted epidemiological health assessment to understand the burden of diseases impacting the PHCC registered population

**Design:** This is a cross-sectional study design among all PHCC registered population between the 1^st^ of September 2018 and the 31^st^ of August 2019

**Setting:** Primary Health Care Corporation health centers

**Participants:** The target population is all persons residing in Qatar aged (0-80) years and registered at the PHCC. Excluding patients with expired Qatar residence permit by the 31^st^ of August

**Results:** Obesity rates ranged between 37% and 35% among the total population registered with the lowest rate in the central region at 34.7%. Burden of type 2 diabetes, hypertension, and dyslipidemia was the highest among population of the Central region at 13.9%, 15.7% and 11.1%, respectively. Tobacco consumption among males ranged from 25.4 % to 27.8%, with the highest rate in the Northern region. 39.9% of females in the Northern region had BMI above 30 kg/m^2^. Exclusive breastfeeding at 6 months was significantly lower than that at 4 months across all regions. Children in the Northern region had the highest rate of overweight/obesity based on Z-scores. Western region population had the highest number of communicable diseases notifications especially Chicken pox at 94.6 per 10,000 children

**Conclusion:** Understanding the patterns of disease in the local population will enable PHCC to provide a clear set of objectives to work towards meeting population health needs

## Introduction

Qatar has a dynamic population. In 2019, it was reported that within the total population of 2,666,938 (August 2019) (1) there were 94 different nationalities residing in Qatar, with 10% of the population being Qatari (2). Qatar’s expat population is fluid. This can make planning for services such as health care challenging. However, much of Qatar’s strategic focus is on the local population, as the long-term residents of the country, and where the greatest impact on health spending and future planning can be made.

Recent demographic and epidemiological trends point to populations living longer and with higher disease burden worldwide (3). Qatar’s Second National Development Strategy (2017-2022) is expected to focus on healthcare, one of eight priority sectors which will be integrated into sector projects, including the delivery of its newest National Health Strategy 2018-2022 which has a strong focus on primary care as the gateway to all other health care services (4). The nationally led programs to establish Integrated Care across the health sector will make great strides in ensuring patients with chronic disease and multiple NCDs will experience a greater coordination of care and better patient outcomes (5).

In Qatar, the highest cause of mortality in both men and women are diseases of the circulatory system (5). Cardiovascular disease is a major cause of health problems and deaths worldwide (6). It is also one of the most preventable causes of death linked to unhealthy diet, lack of exercise, being overweight and smoking. When looking at NCDs collectively, 59.6% of males and 60.7% of females die every year in Qatar from diseases which could be either prevented or better managed to allow patients to live longer and healthier lives (5).

The high rates of NCDs also put considerable strain on secondary care services causing a high rate of hospital attendances and admissions. In addition, the percentage of total diagnoses for patients admitted to HMC in 2018 which was attributable to diabetes was 87% (7). This included diagnoses such as stroke, hypertension, coronary heart disease, chronic obstructive pulmonary disorder and amputation (7). Moreover, the prevalence of type 2 diabetes is projected to grow by 43% and consume one third of Qatar’s health expenditure by 2050 (8).

The Primary Health Care Corporation (PHCC) is the major public sector provider of primary care services to families in Qatar. PHCC operates 27 health centers (March 2020) in three main health regions – Northern, Central and Western. The registered active population in these health centers was 1,461,987 by February 2020 - a 55% increase since 2015 (9).

The epidemiological health assessment will provide a better understanding of PHCC’s targeted population health needs, risk factors, and prevalence of diseases. The data will support better service design, implementation, and resource allocation to respond to the population health needs.

### Aim

The epidemiological health assessment aims at providing a better understanding of the Primary Heath Care Corporation (PHCC) targeted population health needs, risk factors, and prevalence of diseases by the three regions that PHCC health centers operate in. The data will support better service design, implementation, and resource allocation to respond to the population health needs.

### Objectives

- To assess the prevalence of chronic non-communicable diseases and their modifiable behavioral/metabolic risk factors among the population registered at PHCC facilities
- To assess the prevalence of depression and anxiety among the registered population at PHCC health facilities
- To provide up-to-date information for assessing the situation of newborns, children and women registered at primary health care facilities in Qatar.
- To understand the incidence of the communicable diseases reported at the PHCC health centers

## Methodology

### Study design

A cross-sectional study design was implemented where participants were observed only once, and data was collected once. Cross-sectional study design is a type of observational study that analyzes data from the population or representative sample of the population to provide information on the prevalence of diseases or conditions among the studied population.

### Study setting

Primary Health Care Corporation

### Study population

The target population is all persons residing in Qatar aged 0-80 years and registered at the PHCC, a total of 1,247,183 person.

- **Inclusion criteria** All PHCC registered population with a valid Health Card between the 1^st^ of September 2018 and the 31^st^ of August 2019.
- **Exclusion criteria** Registered PHCC patients with expired Qatar residence permit by the 31^st^ of August.

## Data collection and data analysis

Data was extracted electronically from Electronic Medical Records (EMR) covering the period between 1^st^ of September 2018 and 31^st^ of August 2019 per health center. The data extraction included all the available information on the non-communicable diseases (diabetes, hypertension, dyslipidemia, depression and anxiety); metabolic and behavior risk factors (tobacco consumption, BMI, Lipid profile, and Glaucous profile); breastfeeding practices, and the z-score for weigh-for-age captured during the well-baby clinics visits; and the communicable diseases notifications.

Descriptive analysis using STATA 14 was applied to calculate proportion of diseases and condition against their target population.

The burden of non-communicable diseases (diabetes, hypertension, dyslipidemia, depression and anxiety) were assessed by using the diagnosis field excluding gestational diabetes and hypertension divided by population at risk (population aged 18+). Metabolic and behavior risk factors were assessed by using the latest available value captured in the system between 1^st^ of September 2018 and 31^st^ of August 2019. Breastfeeding practices were evaluated based on the mothers’ responses to the breastfeeding embedded questionnaire in Cerner at their visits to the well-baby clinics at 4 months and 6 months of the infant age. The top communicable diseases notifications were reported by their number, their percentage of the total notification, and by their incidence per 10000 to their respective target population age-group and nationality.

Data was presented as per the three regions that the PHCC cluster is operating health centers under to better understand the disparities of burden of diseases in the regions.

## Results

### 1. Demography

The highest number of registered population excluding the deceased and the persons with expired residency permit at the 31st of August 2019 point in time was in the central region at 531,568 persons. Children and young people aged between 0 to 18 years constitute almost 30% of the PHCC registered population across the three regions (table No.1, No.2 and No.3). Female percentage out of the total registered population was 45% in the western region and 48% in central and northern regions (tables No. 1, No.2 and No.3).

**Table No.1:**
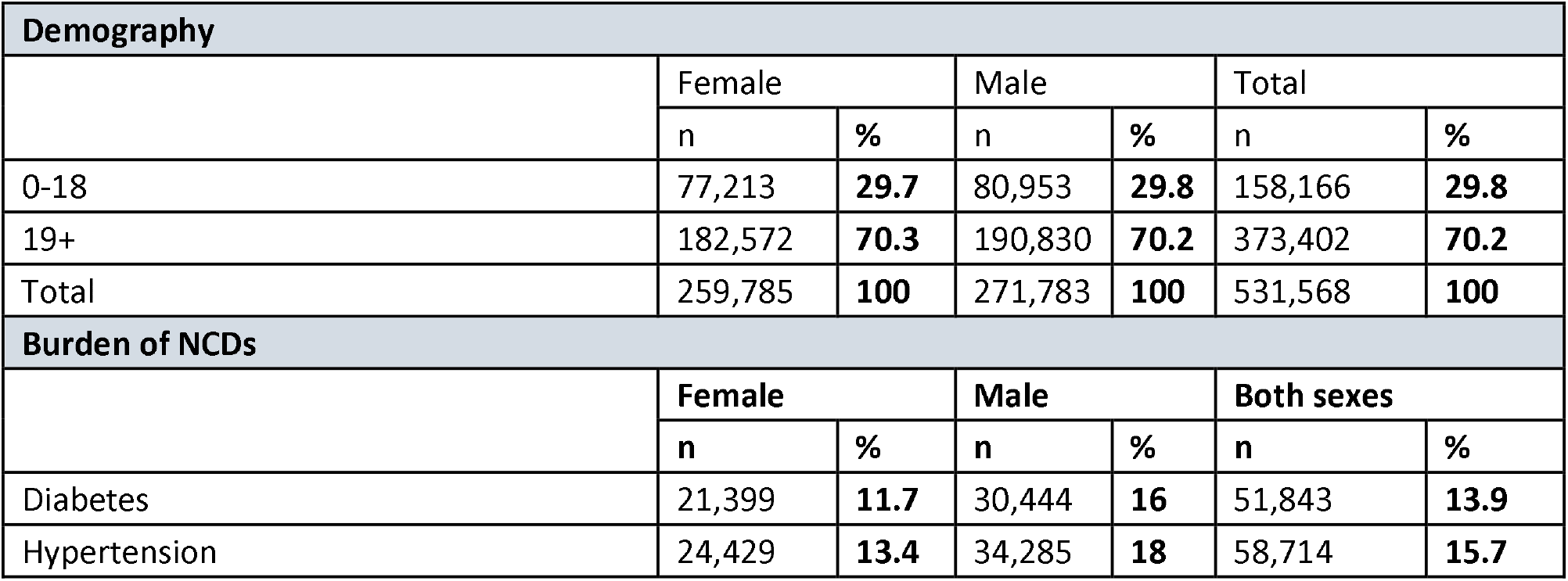
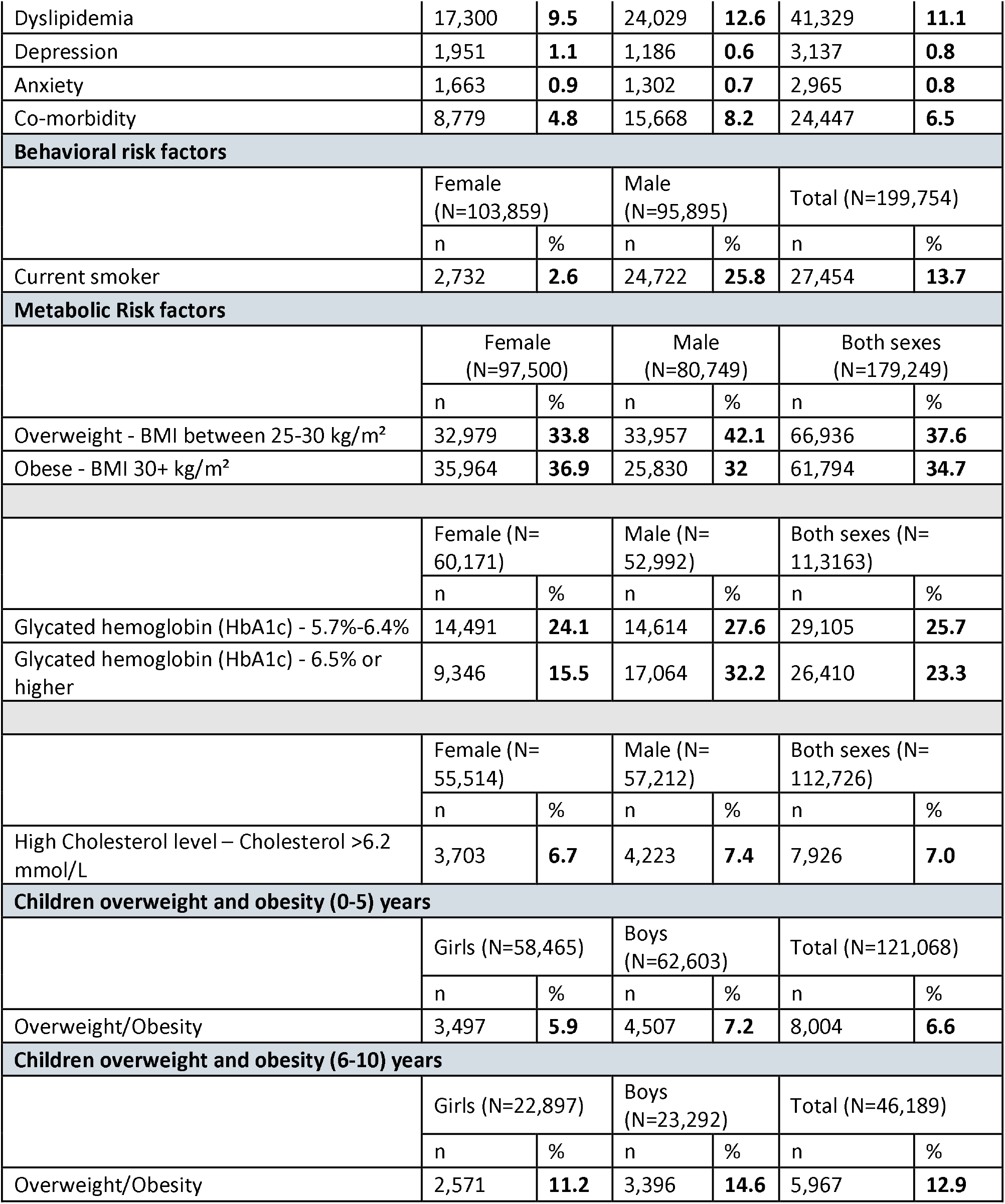
PHCC central region epidemiological health assessment by sex.

**Table No.2:** PHCC western region epidemiological health assessment by sex.

| **Demography** |  |  |  |  |  |  |
| --- | --- | --- | --- | --- | --- | --- |
|  | Female |  | Male |  | Total |  |
|  | n | % | n | % | n | % |
| 0-18 | 53,567 | **34.9** | 56,231 | **30.8** | 109,798 | **32.7** |
| 19+ | 99,812 | **65.1** | 126,625 | **69.2** | 226,437 | **67.3** |
| Total | 153,379 | **100** | 182,856 | **100** | 336,235 | **100** |
| **Burden of NCDs** |  |  |  |  |  |  |
|  | Female |  | Male |  | Both sexes |  |
|  | n | % | n | % | n | % |
| Diabetes | 12,641 | **12.7** | 14,352 | **11.3** | 26,993 | **11.9** |
| Hypertension | 11,776 | **11.8** | 14,982 | **11.8** | 26,758 | **11.8** |
| Dyslipidemia | 7,484 | **7.5** | 10,095 | **8** | 17,579 | **7.8** |
| Depression | 832 | **0.8** | 459 | **0.4** | 1,291 | **0.6** |
| Anxiety | 776 | **0.8** | 665 | **0.5** | 1,441 | **0.6** |
| Co-morbidity | 4,162 | **4.2** | 7,096 | **5.6** | 11,258 | **5.0** |
| **Behavioral risk factors** |  |  |  |  |  |  |
|  | Female<br>(N=52,502) |  | Male<br>(N=53,645) |  | Total (N=106,147) |  |
|  | n | % | n | % | n | % |
| Current smoker | 1201 | **2.3** | 13645 | **25.4** | 14846 | **14.0** |
| **Metabolic risk factors** |  |  |  |  |  |  |
|  | Female<br>(N=59,075) |  | Male<br>(N=44,147) |  | Both sexes<br>(N=103,222) |  |
|  | n | % | n | % | n | % |
| Overweight - BMI between 25-30 kg/m<sup>2</sup> | 19,233 | **32.6** | 17,733 | **40.2** | 36,966 | **35.8** |
| Obese - BMI 30+ kg/m<sup>2</sup> | 23,525 | **39.8** | 14,637 | **33.2** | 38,162 | **37.0** |
|  | Female (N=35,937) |  | Male (N=28,939) |  | Both sexes (N=64,876) |  |
|  | n | % | n | % | n | % |
| Glycated hemoglobin (HbA1c) - 5.7%-6.4% | 8,230 | **22.9** | 7,924 | **27.4** | 16,154 | **24.9** |
| Glycated hemoglobin (HbA1c) - 6.5% or higher | 5,229 | **14.6** | 8,381 | **29** | 13,610 | **21.0** |
|  | Female (N=31,646) |  | Male (N=30,071) |  | Both sexes (N=61,717) |  |
|  | n | % | n | % | n | % |
| High Cholesterol level – Cholesterol >6.2 mmol/L | 1,915 | **6.1** | 2,371 | **7.9** | 4,286 | **6.9** |
| **Children overweight and obesity (0-5) years** |  |  |  |  |  |  |
|  | Girls (N=68,157) |  | Boys (N=73,143) |  | Total (N=141,300) |  |

|  | n | % | n | % | n | % |
| --- | --- | --- | --- | --- | --- | --- |
| Overweight/Obesity | 3,957 | **5.8** | 5132 | **7** | 9,089 | **6.4** |
| **Children overweight and obesity (6-11) years** |  |  |  |  |  |  |
|  | Girls (N=32,285) |  | Boys (N=34,670) |  | Total (N=66,955) |  |
|  | n | % | n | % | n | % |
| Overweight/Obesity | 3,331 | **10.3** | 4,453 | **12.8** | 7,784 | **11.6** |

**Table No.3:** PHCC northern region epidemiological health assessment by sex.

| **Demography** |  |  |  |  |  |  |
| --- | --- | --- | --- | --- | --- | --- |
|  | Female |  | Male |  | Total |  |
|  | n | % | n | % | n | % |
| 0-18 | 57,595 | **31.1** | 60,493 | **31.2** | 118,088 | **31.1** |
| 19+ | 127,781 | **68.9** | 133,511 | **68.8** | 261,292 | **68.9** |
| Total | 185,376 | **100** | 194,004 | **100** | 379,380 | **100** |
| **Burden of NCDs** |  |  |  |  |  |  |
|  | Female |  | Male |  | Both sexes |  |
|  | n | % | n | % | n | % |
| Diabetes | 12,641 | **12.7** | 14,352 | **11.3** | 26,993 | **11.9** |
| Hypertension | 11,776 | **11.8** | 14,982 | **11.8** | 26,758 | **11.8** |
| Dyslipidemia | 7,484 | **7.5** | 10,095 | **8** | 17,579 | **7.8** |
| Depression | 832 | **0.8** | 459 | **0.4** | 1,291 | **0.6** |
| Anxiety | 776 | **0.8** | 665 | **0.5** | 1,441 | **0.6** |
| Co-morbidity | 4,162 | **4.2** | 7,096 | **5.6** | 11,258 | **5** |
| **Behavioral risk factors** |  |  |  |  |  |  |
|  | Female (N=53,190) |  | Male (N=45,014) |  | Total (N=98,204) |  |
|  | n | % | n | % | n | % |
| Current smoker | 1105 | **2.1** | 12513 | **27.8** | 13618 | **13.9** |
| **Metabolic risk factors** |  |  |  |  |  |  |
|  | Female (N=66,295) |  | Male (N=49,738) |  | Both sexes (N=116,033) |  |
|  | n | % | n | % | n | % |
| Overweight - BMI between 25-30 kg/m<sup>2</sup> | 21,253 | **32.1** | 20,270 | **40.8** | 41,523 | **35.8** |
| Obese - BMI 30+ kg/m<sup>2</sup> | 26,433 | **39.9** | 16,039 | **32.2** | 42,472 | **36.6** |
|  | Female (N=42,054) |  | Male (N=33,002) |  | Both sexes (N=75,056) |  |
|  | n | % | n | % | n | % |
| Glycated hemoglobin (HbA1c) - 5.7%-6.4% | 9,702 | **23.1** | 9,041 | **27.4** | 18,743 | **25** |
| Glycated hemoglobin (HbA1c) - 6.5% or higher | 6,534 | **15.5** | 9,601 | **29.1** | 16,135 | **21.5** |

|  | Female (N=37,967) |  | Male (N=34,148) |  | Both sexes (N=72,115) |  |
| --- | --- | --- | --- | --- | --- | --- |
|  | n | % | n | % | n | % |
| High Cholesterol level – Cholesterol >6.2 mmol/L | 2,518 | **6.6** | 2,785 | **8.2** | 5,303 | **7.4** |
| **Children overweight and obesity (0-5) years** |  |  |  |  |  |  |
|  | Girls (N=49,728) |  | Boys (N=53,135) |  | Total (N=102,863) |  |
|  | n | % | n | % | n | % |
| Overweight/Obesity | 3,178 | **6.4** | 4,198 | **7.9** | 7,376 | **7.2** |
| **Children overweight and obesity (6-11) years** |  |  |  |  |  |  |
|  | Girls (N=21,661) |  | Boys (N=22,096) |  | Total (N=43,757) |  |
|  | n | % | n | % | n | % |
| Overweight/Obesity | 2,578 | **11.9** | 3,216 | **14.5** | 5,794 | **13.3** |

### 2. Burden of noncommunicable diseases

Central region registered population appeared to have the highest rates of non-communicable diseases. The burden of type 2 diabetes excluding gestational diabetes was the highest among the PHCC registered population in the central region at 13.9% followed by the population registered at the northern and western regions at 12.6% and 11.9% respectively. The highest rate was among the male population in the Central region at 16%. Hypertension rate excluding gestational was the highest among the registered population in the central region at 15.7% followed by population registered at the northern and western region at 12.5% and 11.8% respectively. Males in the central region had the highest rate at 18%. Burden of dyslipidemia was the highest in the central region at 11.1% while it was the lowest in the western region at 7.8%. Male population registered at the central region had the highest rate at 12.6%. Depression rates ranged between 0.8% and 0.6% among the total population across the three regions with the highest rate identified among females registered at the central region at 1.1%. Co-morbidity of diabetes and hypertension ranged between 6.5% and 5% with the highest rate registered at the central region (tables No. 1, No.2 and No.3).

### 3. NCD risk factors prevalence

Tobacco consumption among males ranged from 25.4 % to 27.8%, with the highest rate in the northern region as dementated in tables No. 1, No.2 and No.3.

Obesity rates ranged between 37% and 35% among the total population registered in the three regions with the lowest rate in the central region at 34.7%. Females had more BMI values above 30 kg/m^2^ than males with the highest rate among the females in the northern region at 39.9% (tables No. 1, No.2 and No.3).

Glycated hemoglobin (HbA1c) - 6.5% or higher was the highest among the population registered at the central region at 23.3 % followed by the population registered at the northern and western region at 21.5% and 21.0%, respectively. The highest rate was among the male population in the central region at 32.2% (tables No. 1, No.2 and No.3.)

High cholesterol level - Cholesterol >6.2 mmol/L was highest among the population registered at the northern region at 7.4%. The rate was similar among the population registered in the central and western region at 7% (tables No. 1, No.2 and No.3).

### 4. Children overweight and obesity

Z (standard) score are used to describe weight-for-age as the number of standard deviations above or below the reference mean/median value. The Z-score is gender independent which permits comparison across populations and different age groups. The prevalence of overweight/obesity was consistently higher among children in the age groups (6–10) years old for both boys and girls when compared to those in the age group (0-5) in all geographical regions. In addition, the prevalence of overweight/obesity was consistently higher among boys when compared to girls regardless of age in all geographical regions. In the Northern region, overweight/obesity among younger children aged (0-5) was the highest at 7.2% compared to 6.6% and 6.4% in Central and Western regions, respectively. The same trend was observed among older children aged (6–10) where those residing the Northern region had the highest prevalence of overweight/obesity at 13.3% compared to 12.9% and 11.6% in Central and Western regions, respectively (tables No. 1, No.2 and No.3)

### 5. Breastfeeding practices

Exclusive breastfeeding at 6 months was significantly lower than that at 4 months. This trend was observed in the three regions. In the Central region, exclusive breastfeeding at 4 months among mothers was the highest at 30.7% compared to 26% and 25.7% in northern and western, respectively. Exclusive breastfeeding at 6 months ranged 7.1% and 9.5% in the three regions (table No.4).

**Table No.4:** Exclusive breastfeeding practices at 4 and 6 months by region.

| **Central region** |  |  |  |  |
| --- | --- | --- | --- | --- |
|  | 4 Months (N=2,815) |  | 6 Months (N=2,906) |  |
| Breast Feeding Practice | n | % | n | % |
| Exclusive breast feeding | 850 | **30.2** | 207 | **7.1** |
| **Western region** |  |  |  |  |
|  | 4 Months (N=2,303) |  | 6 Months (N=2,228) |  |
| Breast Feeding Practice | n | % | n | % |
| Exclusive breast feeding | 591 | **25.7** | 212 | **9.5** |
| **Northern region** |  |  |  |  |
|  | 4 Months (N=1,419) |  | 6 Months (N=1,735) |  |
| Breast feeding practice | n | % | n | % |
| Exclusive breast feeding | 369 | **26.0** | 151 | **8.7** |

### 6. Communicable diseases notifications

Western region registered population had the highest number of communicable diseases notifications. Chicken pox notifications had the highest incidence rate at 94.6 child per 10,000 child aged between 5 to 9 years while in central and northern region the rate ranged from 47.2 and 48.8. Head lice notifications was the highest in the western region at 32.4 per 10,000 child aged between 4 to 5 years. In addition, there was a higher incidence of MERS (Middle East respiratory syndrome) in the Western region at 8.6 per 10,000 persons (table No.5).

**Table No.5:**
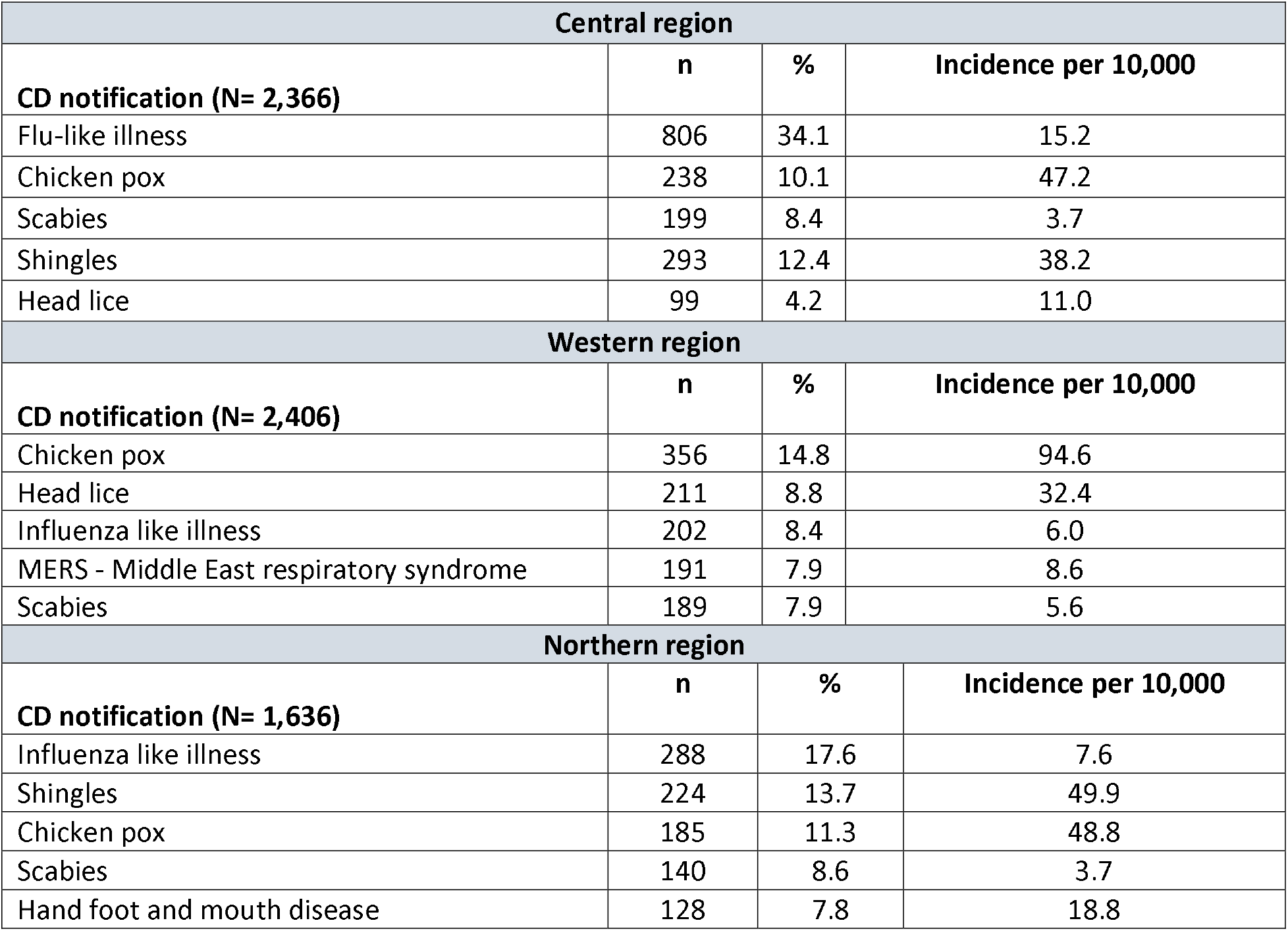
Communicable diseases notification by region.

## Discussion

Our comprehensive review and assessment of data collected through the PHCC’s epidemiological health assessment for its registered population has identified multiple health concerns among PHCC’s registered population.

Non-communicable diseases kill 40 million people each year, equivalent to 70% of all deaths globally (10). Each year, 15 million people die from a chronic non-communicable between the ages of 30 and 69 years; over 80% of these “premature” deaths occur in low- and middle-income countries (10). Modifiable behavioral risk factors such as tobacco use, unhealthy diet and physical inactivity and harmful use of alcohol all increase the risk of NCD. Tobacco is responsible for over 7.2 million deaths every year (11). Metabolic risk factors increase the risk of NCD with raised blood pressure as a main contributor for global mortality (11).

Qatar has undergone tremendous change in the socioeconomic status of its population. This have contributed to change in lifestyle that is characterized by poor nutrition and lack of exercise. All of these mentioned factors have led to an overall increase in the prevalence of obesity, diabetes mellitus, and NCDs in general (12)

In Qatar, the prevalence of diabetes among the Qatari population above 18 years old was 16.7 % and the prevalence of high blood pressure among the same group was 33% (13). Recent estimates show that the prevalence of diabetes mellitus will increase from 16.7% to at least 24.0% by 2050” (8). Our study shows that obesity rates ranged between 37% and 35% among the total population registered in the three geographical regions of PHCC. This is similar to results outlined earlier in Qatar’s STEPS report conducted in 2012 that reported an obesity prevalence of 40% among Qataris aged 18+ years (13). Al Thani M and colleagues reported an overall overweight and obesity prevalence of 44.8% and 40.4%, respectively, among students in Qatar. In addition, they found that among students “males had 1.48 times higher odds of having obesity than females” (12). We noticed the same trend in our study that demonstrated that among children aged (0-10) the prevalence of overweight/obesity was consistently higher among boys when compared to girls in all geographical regions. We also found that the rate of obesity increased with the increase of age among those children.

Tobacco consumption among males ranged from 25.4 % to 27.8% among our study population, which is higher than the overall smoking prevalence of 16.4% reported by Haj Bakri A et al in 2012 in Qatar (13).

A recent study conducted on primary care population in Qatar had found that around 16.2% of the studied population had at least one of the following NCDs: type 2 diabetes, cardiovascular diseases, chronic obstructive pulmonary diseases, or cancer. In addition, they reported that the burden of NCDs was higher among men when compared to women (14). Our study supports those findings as it shows that the highest rates of type 2 diabetes, excluding gestational diabetes, Hypertension, and dyslipidemia were among the male population in the Central region at 16%, 18%, and 12.6% respectively.

The importance of this study comes from providing updated figures on the situation of NCDs and their subsequent risk factors in Qatar among all the registered population at the PHCC and by stratifying the results per region to assess and to understand whether the geographical location in Qatar has an effect on the disease and risk pattern. Therefore, and with this study, there was a trend observed for the NCDs having the highest prevalence among the population registered in the central region. A possible explanation for that could be because the Central region includes the main big cities in Qatar namely: Doha, and Al Wakra where people have higher awareness about disease and better access to screening and diagnosis. However, this needs to be explored more in future research. The highest prevalence for NCD risk factors was observed among the population in the northern region.

The Western region population had the highest number of communicable diseases notifications especially chicken pox at 94.6 child per 10,000 child aged between 5 to 9 years. Although “Qatar was the first in the Middle East region to introduce the chickenpox vaccine (marketed as Varilrix®) in 2002” (15), we still see varicella infections. Chickenpox is more prevalent among school-age children compared to younger children. Historical vaccination data collected at the primary care level in Qatar suggests that these children might have missed their second vaccine dose which aims to boost the immunity of the children and enhance the protective effect of the vaccine. In addition, the long timespan between the first dose administered at 12 months and second one administered between 4 to 6 years is a contributing factor for vaccine default. Finally, we need to keep in mind that varicella vaccine is not routine in many countries, including countries of origin for vast majority of non-Qatari population. It’s important to have continuous surveillance to examine the need for catchup immunizations especially for populations at risk that have interrupted or delayed vaccinations (15).

Breastfeeding practices still remain low in Qatar; the available data from 2012 showed that only 29.3% percent of infant aged 0-5 months were exclusively breastfed in Qatar (16). Another recent study conducted by the University of Qatar in 2017 reported that the total number of children being exclusively breastfed (12%) is significantly lower than the international rate (37%) (17). And, it identified some of the barriers to continued breastfeeding and they include perception of breastfeeding as painful, body image, difficulty of breastfeeding in public, or at work (17).

In our study, exclusive breastfeeding at 6 months was significantly lower than that at 4 months. This trend was observed in the three regions. In the central region, exclusive breastfeeding at 4 months among mothers was the highest at 30.7% compared to 26% and 25.7% in the northern and western regions, respectively. These findings highlight the crucial need for community-based programs that encourages exclusive and continued breastfeeding. Health educators and other health care providers can work with the mothers to support them, improve their breastfeeding skills and engage other family members to provide additional support for them. In addition, this needs to be communicated with policy makers to advocate for workplaces that are breastfeeding friendly and for longer maternity leave (17).

## Conclusion

By understanding the patterns of disease in the local population, the PHCC will be able to provide a clear set of objectives to work towards improving the health conditions of its target population. The importance of assessing health needs rather than reacting to health demands is widely recognized. The above findings will be integrated into the planning and commissioning of PHCC’s health services across the country. Recommendations include the increased use of telemedicine services, enhanced education regarding the adverse health effects of tobacco consumption and implementing additional measures to encourage healthy eating habits and exercise, to reduce the prevalence of type 2 diabetes, obesity, and hypertension.

## Strengths and limitations

The main limitation of the study was that the data was collected retrospectively through the patients’ electronic medical records with the latest available value captured in the system during the defined period of time. The latter might subject the study to incomplete data for certain variables, namely the social behavior variables.

The strength of the epidemiological health assessment derives from including all the population registered at the PHCC between the 1st of September 2018 and 31st of August 2019. The cross section study design allowed to have data on the whole study population at a single point in time. The latter allowed to assess and understand the frequencies and the distribution of the NCDs and their subsequent risk factors, breastfeeding practices, and communicable diseases notifications across the PHCC target population by sex and region of registration.

## Data Availability

The data is available upone request.

## Foot notes

### Ethics approval

The epidemiological health assessment was approved by the PHCC Corporate Strategy Implementation group at PHCC in April 2019. The project was exempt from IRB approval as data was extracted retrospectively as part of epidemiological activities to support better development of the PHCC key strategic activities for better service provision towards its target population. The project was presented later at the PHCC scientific research committee representing the PHCC IRB committee in which it approved the study and provided the required ethical approval for conducting this study.

### Provenance and peer review

Not commissioned; externally peer reviewed.

### Data availability statement

No additional data are available.

